# Mapping SARS-CoV-2 Antibody Epitopes in COVID-19 Patients with a Multi-Coronavirus Protein Microarray

**DOI:** 10.1101/2021.01.14.21249690

**Authors:** David Camerini, Arlo Z. Randall, Krista Trappl-Kimmons, Amit Oberai, Christopher Hung, Joshua Edgar, Adam Shandling, Vu Huynh, Andy A. Teng, Gary Hermanson, Jozelyn V. Pablo, Megan M. Stumpf, Sandra N. Lester, Jennifer Harcourt, Azaibi Tamin, Mohammed Rasheed, Natalie J. Thornburg, Panayampalli S. Satheshkumar, Xiaowu Liang, Richard B. Kennedy, Angela Yee, Michael Townsend, Joseph J. Campo

## Abstract

The emergence and rapid worldwide spread of SARS-CoV-2 has accelerated research and development for controlling the pandemic. A multi-coronavirus protein microarray was created containing full-length proteins, overlapping protein fragments of varying lengths and peptide libraries from SARS-CoV-2 and four other human coronaviruses. Sera from confirmed COVID-19 patients as well as unexposed individuals were applied to multi-coronavirus arrays to identify specific antibody reactivity. High level IgG, IgM and IgA reactivity to structural proteins S, M and N, as well as accessory proteins, of SARS-CoV-2 were observed that was specific to COVID-19 patients. Overlapping 100, 50 and 30 amino acid fragments of SARS-CoV-2 proteins identified antigenic regions. Numerous proteins of SARS-CoV, MERS-CoV and the endemic human coronaviruses, HCoV-NL63 and HCoV-OC43 were also more reactive with IgG, IgM and IgA in COVID-19 patient sera than in unexposed control sera, providing further evidence of immunologic cross-reactivity between these viruses. The multi-coronavirus protein microarray is a useful tool for mapping antibody reactivity in COVID-19 patients.

## Introduction

A novel human coronavirus, which causes severe acute respiratory syndrome, now known as SARS-CoV-2 emerged in December 2019. Infection with SARS-CoV-2 spread rapidly worldwide and on March 11^th^, 2020, it was declared a pandemic by the World Health Organization (WHO; 1). As of November 30^th,^ 2020, there are over 63 million confirmed cases of coronavirus disease (COVID-19) caused by this new virus, resulting in more than 1.4 million deaths, corresponding to a case mortality rate of ∼2.3% (2). Best current estimates indicate that SARS-CoV-2 has a basic reproductive number, R_0_, of 2 to 2.5 and an incubation time of approximately 4.6 days (3, 4), which allows rapid spread of the virus. Diagnosis, treatment and vaccination against COVID-19 will all benefit from a clear understanding of the immune response to SARS-CoV-2 infection.

Previous studies have shown that COVID-19 patients rapidly seroconvert to SARS-CoV-2 and produce IgM, IgG and IgA antibodies directed to several viral proteins (5-8). Reinfection challenge studies in rhesus macaques showed that the humoral and cellular immune response to SARS-CoV-2 infection was effective in blocking reinfection (25, 26). Nevertheless, it is not clear whether all antibody responses are beneficial or whether some antibody responses to SARS-CoV-2 lead to a less favorable course of disease (9, 10). Moreover, enhancement of infection by antibodies has been reported for severe acute respiratory syndrome coronavirus (SARS-CoV), which is closely related to SARS-CoV-2 (11-13, 24).

We have created and used a multi-coronavirus protein microarray, containing over one-thousand coronavirus proteins, protein fragments and peptides to map IgG, IgA and IgM antibody epitopes in sera from COVID-19 patients. Our approach localizes the antibody reactivity of COVID-19 patients within SARS-CoV-2 proteins and allows us to map the antigenic regions bound. Furthermore, we can similarly measure the antibody reactivity of COVID-19 patients and healthy controls with endemic human coronaviruses and with the two previous epidemic coronaviruses, SARS-CoV and Middle East Respiratory Syndrome coronavirus (MERS-CoV). Our findings and the multi-coronavirus protein microarray we created will be useful in discerning which *de novo* and cross-reactive antibody responses to SARS-CoV-2 are protective and which may be less useful in preventing disease or may even be detrimental. In addition, if high levels of antibody to specific epitopes are found to be especially protective, the array could be used to screen convalescent plasma for therapeutic potential and vaccine recipient sera as a preliminary measure of efficacy (27-30).

## Results

The multi-coronavirus protein microarray created and used in this study encompasses over nine-hundred features. It includes the four structural proteins and five accessory proteins of SARS-CoV-2 as well as overlapping 100, 50 and 30 amino acid (aa) protein fragments of these nine SARS-CoV-2 proteins. It also contains the structural proteins of SARS-CoV, MERS-CoV, HCoV-NL63 and HCoV-OC43, plus overlapping 13-20 aa peptides of the SARS-CoV structural proteins and of the S proteins of MERS-CoV, HCoV-NL63 and HCoV-OC43 (Table 1).

**Table 1.** Features of the 1^st^ Generation ADI Multi-Coronavirus Protein Microarray.

| Virus | S-protein | E-protein | M-protein | N-protein | Accessory or Other Proteins | Total Number of Features = 934 |
| --- | --- | --- | --- | --- | --- | --- |
| SARS-CoV-2 | RBD*, S<sup>#</sup> from BEI, S1, S2, 30, 50 & 100 aa fragments by IVTT at ADI | Protein and 30, 50 aa fragments by IVTT at ADI | Protein and 30, 50, 100 aa fragments by IVTT at ADI | Protein BEI, 30, 50 & 100 aa fragments by IVTT at ADI | ORF 3a, 6, 7a, 8 and 10 protein, 30, 50 & 100 aa fragments by IVTT at ADI | 12 proteins, 310 fragments = 322 features |
| SARS-CoV | Protein** and peptide set from BEI Resources | IVTT by ADI, peptides from BEI | Protein, peptides from BEI | Protein, peptides from BEI | 3CL protease | 5 proteins, 265 peptides = 270 features |
| MERS-CoV | BEI Resources | IVTT by ADI | IVTT by ADI | BEI Resources | ORF 3a, 4a, 4b, 5 and 8b protein IVTT by ADI | 9 proteins |
| HCoV-NL63 | Protein by IVTT at ADI, peptides from BEI | IVTT by ADI | IVTT by ADI | IVTT by ADI | ORF 3 protein IVTT by ADI | 5 proteins and 96 peptides = 101 features |
| HCoV-OC43 | Protein by IVTT at ADI, peptides from BEI | IVTT by ADI | IVTT by ADI | IVTT by ADI | HE, N2 protein IVTT by ADI | 6 proteins and 226 peptides = 232 features |

IVTT means coupled *in vitro* transcription and translation. \* RBD is the receptor binding domain, aa 319 to 541 of the SARS-CoV-2 S protein. # SARS-CoV-2 S protein is a stabilized form with a trimerization sequence and transmembrane domain deletion. \*\* SARS-CoV-S protein is a transmembrane domain deleted form.

The multi-coronavirus array was incubated with sera from two sets of patient samples and associated negative controls collected in different regions of the USA. The first set of sera from ten COVID-19 patients and ten pre-pandemic healthy donors was obtained from the Centers for Disease Control and Prevention (CDC) in Atlanta, Georgia. The second set included sera from ten COVID-19 patients and nine pre-pandemic samples obtained from the Mayo Clinic in Rochester, Minnesota. The age, sex and SARS-CoV-2 ELISA result of the COVID-19 patient and healthy control blood donors in both sample sets is shown in Tables 2 and 3. ELISA’s were performed separately at the two different sites and discriminated the COVID-19 patient samples from the control samples.

**Table 2.** Serum Donors for Samples Obtained from the CDC.

| Sample ID | SARS-CoV-2 S ELISA (S/T) | Age Decade | Sex |
| --- | --- | --- | --- |
| C1 | 0.25 | 60's | M |
| C2 | 0.09 | 20's | M |
| C3 | 0.18 | 40's | M |
| C4 | 0.34 | 50's | M |
| C5 | 0.19 | 60's | M |
| C6 | 0.94 | 40's | M |
| C7 | 0.26 | 40's | M |
| C8 | 0.15 | 50's | M |
| C9 | 0.40 | 40's | M |
| C10 | 0.19 | 50's | M |
| P1 | 2.06 | 60's | F |
| P2 | 3.98 | 60's | M |
| P3 | 5.47 | 50's | F |
| P4 | 2.35 | 20's | F |
| P4 | 5.16 | 70's | F |
| P6 | 5.21 | 70's | M |
| P7 | 3.82 | 30's | M |
| P8 | 5.30 | 80's | M |
| P9 | 3.81 | 40's | F |
| P10 | 5.20 | 40's | M |

S/T = signal/threshold for positivity ratio; >1.0 is positive
C = healthy negative control; P = COVID-19 patient

**Table 3.**
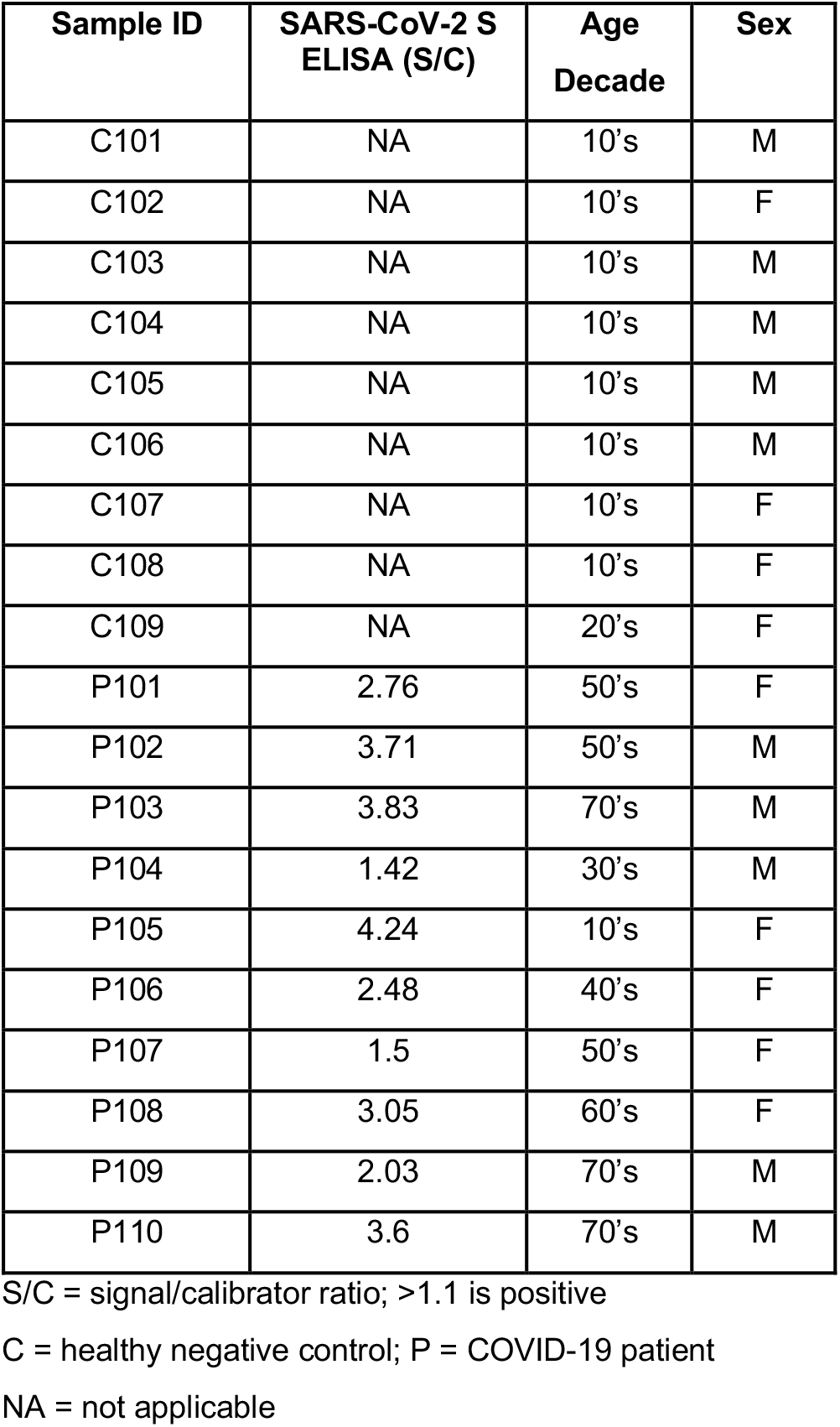
Serum Donors for Samples Obtained from the Mayo Clinic.

| Sample ID | SARS-CoV-2 S ELISA (S/C) | Age Decade | Sex |
| --- | --- | --- | --- |
| C101 | NA | 10's | M |
| C102 | NA | 10's | F |
| C103 | NA | 10's | M |
| C104 | NA | 10's | M |
| C105 | NA | 10's | M |
| C106 | NA | 10's | M |
| C107 | NA | 10's | F |
| C108 | NA | 10's | F |
| C109 | NA | 20's | F |
| P101 | 2.76 | 50's | F |
| P102 | 3.71 | 50's | M |
| P103 | 3.83 | 70's | M |
| P104 | 1.42 | 30's | M |
| P105 | 4.24 | 10's | F |
| P106 | 2.48 | 40's | F |
| P107 | 1.5 | 50's | F |
| P108 | 3.05 | 60's | F |
| P109 | 2.03 | 70's | M |
| P110 | 3.6 | 70's | M |

S/C = signal/calibrator ratio; >1.1 is positive
C = healthy negative control; P = COVID-19 patient
NA = not applicable

### Specific antibody reactivity to SARS-CoV-2 and SARS-CoV purified recombinant proteins in COVID-19 patients

The specimens from COVID-19 patients had robust anti-SARS-CoV-2 IgG and IgA antibodies. IgM antibody responses were weaker. The magnitude and specificity of the antibody responses were similar in both sets of samples, so they are presented together here. COVID-19 patient serum IgG, IgA and IgM reacted strongly to purified SARS-CoV-2 spike (S), as well as SARS-CoV nucleocapsid (N), S and membrane (M) proteins compared to healthy control sera (Fig. 1). The receptor binding domain of the SARS-CoV-2 S protein (RBD) had overall weaker antibody binding signals, but nevertheless was significantly more reactive in COVID-19 patient serum IgG and IgA. The signals shown in figure 1 are the base two logarithm of the raw intensities without normalization, since the background reactivity of each purified protein is different. The SARS-CoV-2 S and RBD as well as the SARS-CoV S, N and M purified proteins had the largest mean differences between IgG binding of the negative and positive groups, and the differences are the most statistically significant (t-test *p* values <10^−5^). The same five antigens had the largest significant mean differences between IgA binding of the negative and positive groups. Only SARS-CoV-2 S and SARS-CoV N, however, had significant differential IgM binding between the COVID-19 patients and the control group. These results are in agreement with the enzyme linked immunosorbent assays (ELISA) shown in Tables 2 and 3.

**Figure 1.**
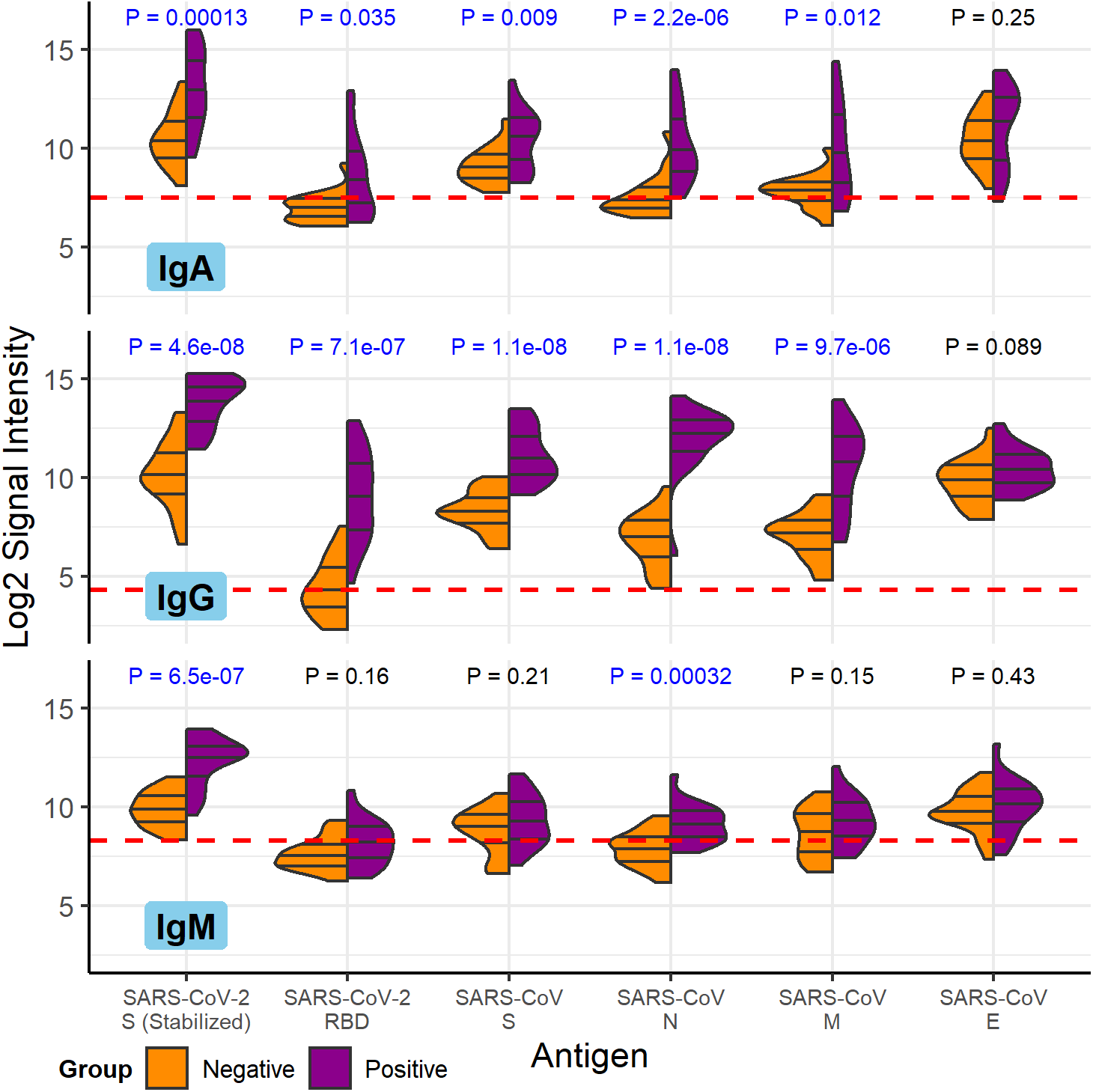
COVID-19 patient and healthy control antibody reactivity with purified SARS-CoV-2 and SARS-CoV proteins. The split violin plot shows the log_2_-transformed fluorescence signal intensity distribution of antibodies bound to each purified protein on the multi-coronavirus protein microarray. Within each half-violin are three lines representing the interquartile range and the median. Above each split violin is the Wilcoxon rank sum *p* value, colored blue for significant *p* values below 0.05. The three panels are split by isotype (IgG, top; IgA, middle; IgM, bottom). Horizontal red dashed lines are drawn at the median of all signal intensities against purified proteins (n=14) and peptides (n=587) plus 1.0, i.e. double the global median—this threshold serves as a point of reference, but not necessarily a seropositivity cutoff for each protein.

### SARS-CoV-2 protein fragments identify antigenic regions

Nine SARS-CoV-2 full-length proteins were produced by coupled *in vitro* transcription and translation (IVTT): S, envelope (E), M, N, open reading frames (ORF’s) 3a, 6, 7a, 8 and 10. We used the same technique to produce overlapping 100 amino acid (aa), 50 aa and 30 aa fragments of each of these nine SARS-CoV-2 proteins and to produce the structural proteins and some accessory proteins of HCoV-NL63, HCoV-OC43 and MERS-CoV. Using aa start and end positions of each fragment within the protein, differential reactivity between the COVID-19 and healthy donor groups was mapped in a circular heatmap for the structural proteins (Fig. 2). This analysis allowed us to identify antigenic regions in each SARS-CoV-2 structural protein. The SARS-CoV-2 N protein showed the strongest reactivity in its carboxy terminal 100 aa fragment, as well as in 50 aa fragments covering the same region. This region was recognized by IgG, IgA and IgM with significant differential reactivity between COVID-19 patients and the healthy control group. The middle of the N protein also had a region recognized by IgG and IgA identified by two 100 aa fragments. Together these antibody-reactive regions encompass about two thirds of the N protein that likely contains at least two epitopes. The S1 protein also showed greatest IgG binding near its carboxy terminus, in the penultimate 100 aa fragment. This antigenic region of S1 was defined further by IgG and IgA reactivity with 50 aa fragments from aa 550 to 600. The region containing the RBD was not strongly reactive when produced by IVTT. In contrast, the S2 protein of SARS-CoV-2 showed three regions of strong IgG, IgA and IgM binding and differential reactivity with full-length, 100 aa and 50 aa fragments. Only the region near the carboxy terminus, however, was also reactive as a 30 aa fragment. This reactive 30 aa fragment, from aa 450 to 480 of S2 (1,135 to 1,165 of S), therefore likely defines a linear IgG epitope in this highly antigenic protein. Notably, an epitope in the central S2 antigenic region was differentially reactive for IgG and IgA, but showed equal levels of IgM reactivity in 100 aa and 50 aa fragments, perhaps indicating a region of cross-reactivity for IgM produced by memory B cells. An additional short epitope was found in the amino terminal 30 aa fragment of the SARS-CoV-2 M protein. This short fragment was highly reactive with COVID-19 patient serum IgG compared to healthy donor serum IgG, while larger fragments containing it and indeed the full-length M protein were not as highly discriminatory for COVID-19 patient sera. The SARS-CoV-2 E protein had only one 30 aa fragment that showed low-level reactivity with IgA and IgM, in both COVID-19 positive and negative control groups. The antigenic regions of SARS-CoV-2 structural proteins we identified were not associated with higher or lower levels of homology between SARS-CoV-2 and other human coronaviruses (percent aa sequence identity shown in outer track of Fig. 2). There was a moderate to high level of correlation between antibody reactivity with S2, N and M proteins produced *in vitro*, particularly for IgG (Pearson’s correlation coefficient shown in inner links of Fig. 2). Less reactivity was seen in non-structural proteins, but significant reactivity of COVID-19 patient sera compared to control sera could be identified in fragments of the 3a and 7a accessory proteins (Supplemental Figure 1).

**Figure 2.**
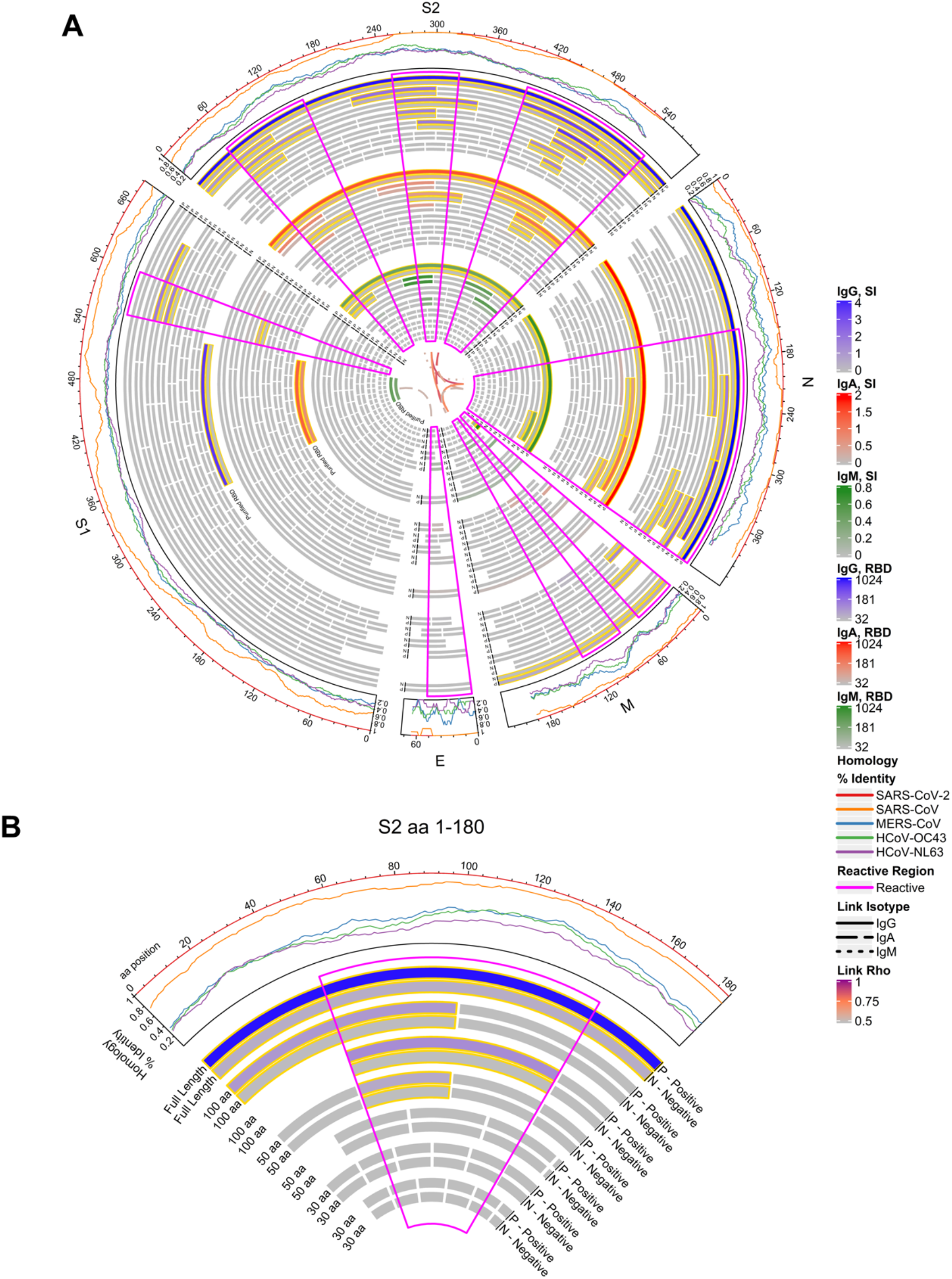
Reactivity of COVID-19 patient and healthy donor IgG (outer band of bars), IgA (middle band) and IgM (inner band) to SARS-CoV-2 proteins and protein fragments. **(A)** The circular graphic maps the amino acid (aa) position of SARS-CoV-2 fragments, showing a heat map of antibody levels in each group for overlapping regions of different aa length. Proteins are indicated outside the circle plot followed by a line graph showing the sequence homology of other CoVs with SARS-CoV-2 for each gene. Proteins and protein fragments produced *in vitro* are indicated by bars and show length and position of each fragment in each protein. Each fragment is drawn twice and shows group mean normalized signal intensity of antibody binding to each fragment for COVID-19 patient samples (P) and negative control sera (N). The purified receptor binding domain (RBD) is additionally shown for comparison. Signal intensity is shown by color gradients: IgG (grey to blue), IgA (grey to red), and IgM (grey to green). Bar pairs shown with gold outline represent significantly differential antibody binding between COVID-19 patients and healthy controls, defined as a mean signal intensity ≥ 0.1 in at least one group and a t-test *p* value ≤ 0.05. The regions of greatest reactivity for each protein are outlined in magenta. Some fragments in E and M proteins that meet the reactivity threshold (grey) and are better visualized by individual responses as shown in Fig. S2. The Pearson’s correlation coefficients (“Rho”) between each full-length protein for each isotype are shown as links between protein sectors in the center of the circle (IgG: solid links, IgA: dashed, IgM: dotted). **(B)** A slice of the circular graphic is amplified and labeled in more detail as a guide to interpreting the full figure. The first 180 aa sequence of S2 is shown for IgG only.

### Individual antibody response profiles to antigenic regions of SARS-CoV-2 and other human coronaviruses

Individual responses to the antigenic regions of SARS-CoV-2 proteins identified by reactivity with protein fragments varied substantially, as they did for the structural proteins of other human coronaviruses (Fig. 3). Responses against the SARS-CoV-2 S1, S2 and N protein fragments, as well as the 30aa fragments of the M protein are shown in Supplemental Figure 2. Within the antigenic regions, some fragments, particularly 30 aa fragments, were nonreactive with COVID-19 patient sera, but others were reactive in a subset of individuals. Heterogeneity was higher and overall signal intensities were lower for IgA and IgM than for IgG. There were no significant associations between age and sex with antibody levels in the positive group after adjustment for the false discovery rate for any of the three isotypes (Supplemental Table 1).

**Figure 3.**
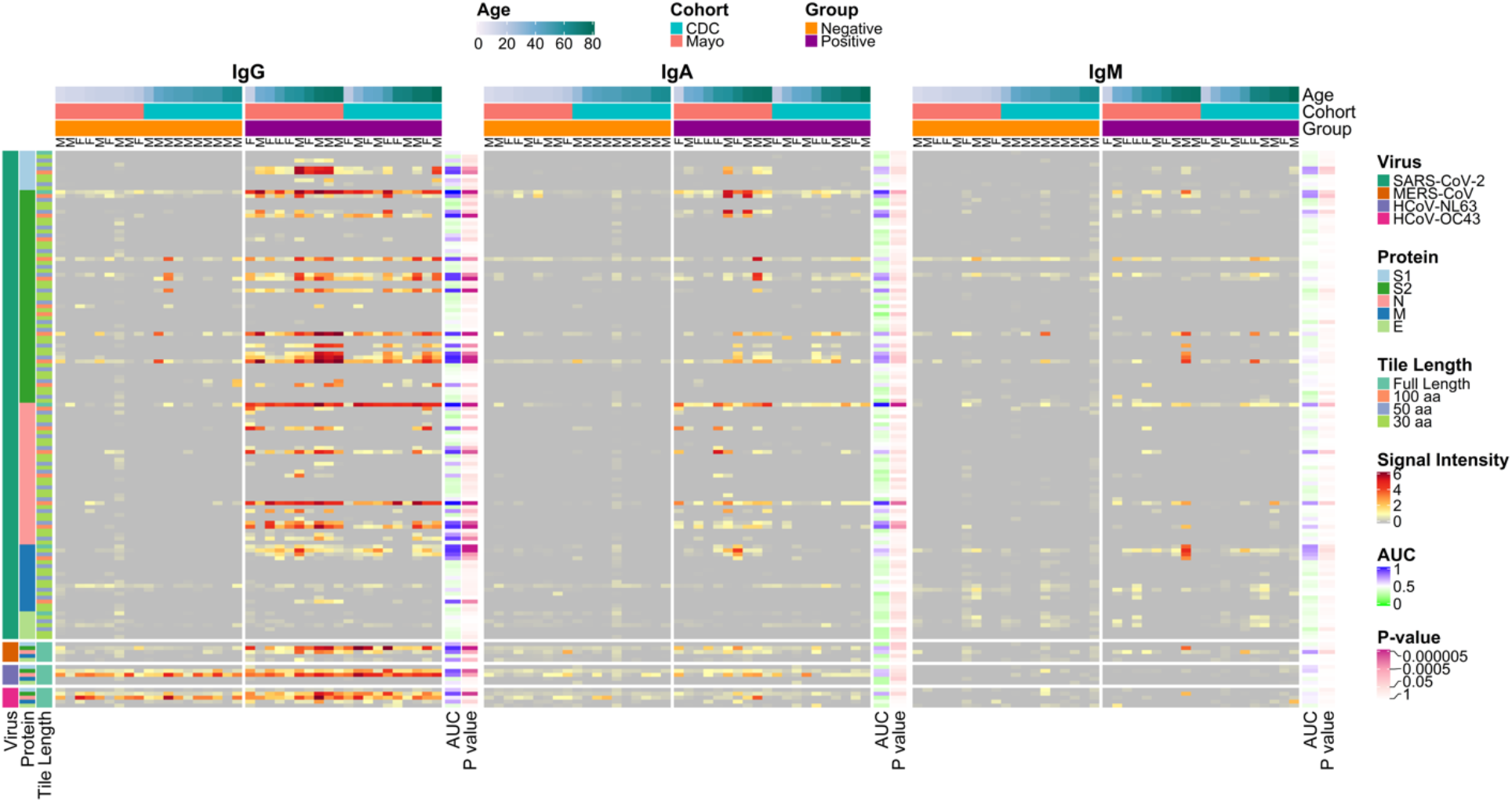
Reactivity in COVID-19 positive and negative IgG (left heatmap), IgA (middle heatmap) and IgM (right heatmap) to SARS-CoV-2 and other HCoV proteins and protein fragments produced *in vitro*. The heatmaps present the signals of antibody binding to individual proteins and protein fragments within the antigenic regions of SARS-CoV-2, as well as the full-length structural proteins of MERS-CoV, HCoV-NL63 and HCoV-OC43, for individual samples. Columns represent serum samples ordered by increasing age within group and cohort, and rows represent proteins or protein fragments; 128 SARS-CoV-2 proteins or fragments, five proteins each of MERS-CoV, HCoV-OC43 and HCoV-NL63. Antibody signal intensity is shown on a color scale from grey to red. Sample information is overlaid above the heatmaps and includes sex (M/F), group (Negative or Positive), cohort (CDC or Mayo) and age (years). Protein/fragment information is annotated to the left of the heatmaps and includes the virus, full-length protein name and the amino acid length of the protein fragments (“Tile Length”, as full length, 100, 50 or 30 aa). For each isotype, the receiver operating characteristic area under the curve (AUC) and the unadjusted t-test *p* value between negatives and positives are shown to the right of each heatmap.

Full-length N proteins of the endemic HCoV’s were reactive with IgG from COVID-19 patients and healthy controls. HCoV-NL63 N was significantly reactive (normalized signal intensity ≥ 1.0) with IgG controls and COVID-19 patients (17/19 and 20/20, respectively; proportions test *p* value = 0.4) (Fig. 3), while HCoV-OC43 N was significantly reactive with IgG control sera and patient sera (15/19 and 20/20, respectively; *p*=0.1). In contrast, IgG from only two control subjects reacted with the SARS-CoV-2 full length N protein while nearly all of the patients’ serum IgG reacted (19/20; *p*=6.8e-7). Reactivity of the control serum IgG with fragments of the SARS-CoV-2 N protein occurred exclusively in the C-terminal region of the protein (1/19) while COVID-19 patient serum IgG reacted frequently with fragments in the central region (12/20; *p*=2.1e-4) of the protein as well as the C-terminal region (19/20; *p*=1.3e-7).

The S2 protein was reactive with patient IgG at a much higher frequency than in the controls for both HCoV-NL63 (5/19 and 16/20 positives, respectively; *p*=2.4e-3) and HCoV-OC43 (4/19 and 18/20, respectively; *p*=5.9e-5). The higher frequencies in the positives provide strong evidence of increased responses due to their exposure to SARS-CoV-2. Some control subject’s IgG reacted with the C-terminal (4/19) or central regions (4/19) of the SARS-CoV-2 S2 protein but none reacted with the N-terminal region; this includes one individual which had unique reactivity to SARS-CoV-2 S2 fragments 401-500 and 451-550 (Figure S2, G). By ELISA, this serum had a S/C value of 0.94, which was just below the positivity threshold of 1.0 and much higher than other healthy donor sera. This reactivity was unique among healthy donors but did not directly translate to reactivity with OC43 or NL63 FL S proteins. Overall, COVID-19 patient serum IgG reacted with the SARS-CoV-2 S2 protein C-terminal (19/20; *p*=1.3e-5), central (17/20; *p*=2.3e-4), and/or N-terminal (12/20; *p*=2.1e-4) 100 aa fragments much more frequently than healthy donor sera.

The reactivity of COVID-19 patient serum IgA compared to IgA of healthy donor sera was similar to results obtained for IgG. The IgA results had lower statistical significance than the IgG results, however, likely due to the lower concentration of IgA in serum compared to IgG. Nevertheless, many of the same proteins were the most differentially reactive with COVID-19 patient serum IgA compared to healthy donor serum IgA, including the N and S proteins and RBD of SARS-CoV-2 as well as the N, S and M proteins of SARS-CoV with t-test *p* values ranging from 2.1 x 10^−6^ to 1.1 x 10^−3^ (Fig. 3). The COVID-19 patient sera used in this study had less coronavirus reactive IgM than IgG or IgA, perhaps because the samples were obtained during the convalescent phase of disease. Nevertheless, significantly greater IgM reactivity was seen in patient sera compared to control donor sera for four proteins and two protein fragments produced *in vitro* (Fig. 3). These were the N, S2 and M proteins of SARS-CoV-2, the MERS-CoV N protein, the carboxy terminal 100 aa fragment of the SARS-CoV-2 N protein and the amino terminal 30 aa fragment of the SARS-CoV-2 M protein.

A library of 587 peptides, 15 to 20 aa in length, from the epidemic SARS-CoV (covering S, N, M and E proteins) and two endemic human coronaviruses (covering S protein) was printed on the multi-coronavirus microarray at the same concentration as full-length purified recombinant proteins. The peptides, however, showed lower antibody reactivity than full length proteins or protein fragments of 30, 50 or 100 aa. Exceptionally, a single 17 aa peptide from HCoV-OC43 S protein with sequence CSKASSRSAIEDLLFDK spanning residues 905 to 921 had approximately 3.5-fold higher mean reactivity in the COVID-19 patient sera (*p*=0.001, not significant after adjustment for the false discovery rate). This peptide mapped to the SARS-CoV-2 sequence PSKPSKRSFIEDLLFNK at residues 809 to 825 of S protein with identical residues in 12/17 positions.

To visualize the relative importance of antibody isotype binding in differentiating COVID-19 positive sera from negative sera, the samples were projected in two dimensions for each isotype using t-distributed stochastic neighbor embedding (tSNE; Fig. 4A), a nonlinear machine learning dimensionality reduction method which clusters together similar sets of multidimensional data. The thirty most reactive proteins for all isotypes were selected for this analysis to reduce the effect of differing isotype background levels that would be notable in low-reactivity proteins (Fig. 4B). Each of the isotypes cluster separately, but only IgG gave a clear delineation of positives and negatives (at ∼2.6 in tSNE dimension 2). Henceforth, the focus of the presented data is on the IgG responses.

**Figure 4.**
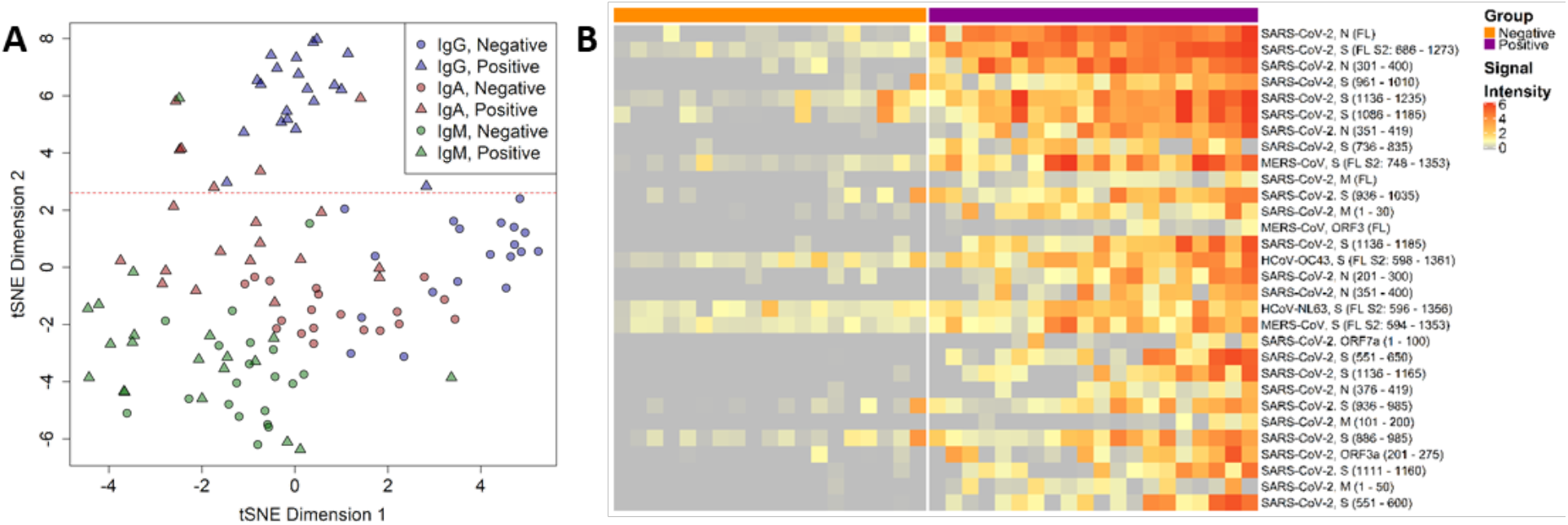
IgG responses give the best delineation of COVID-19 patient sera from healthy donor sera. **(A)** The IgG, IgA and IgM responses against the 30 most reactive IVTT proteins by mean of all samples and isotypes were projected for each sample across two dimensions using t-distributed stochastic neighbor embedding (tSNE). Points represent individual samples and are colored according to the isotype measurement. The shape represents the group in which the sample belonged, either the “Negative” healthy donor group or the “Positive” COVID-19 patient group. The horizontal red dashed line (y=2.6) separates the IgG responses in negative and positive individuals. **(B)** The heatmap shows the 30 most differentially reactive IgG responses to IVTT proteins between the negative and positive groups. Columns represent serum samples, separated by group with colored headers. Rows represent full-length or fragmented proteins produced by cell-free expression *in vitro*. The protein annotations to the right of the heatmap denote the virus, protein and in parentheses the amino acid range of the fragment or full length “FL” protein. Normalized signal intensity is displayed on a grey to red color scale.

The full-length SARS-CoV-2 N and S2 proteins as well as several fragments of both proteins had the top nine largest mean differences in IgG reactivity between COVID-19 patients and healthy controls (Fig. 4B). These results were also statistically significant with t-test *p* values ranging from 2.1 x 10^−6^ to 4.3 x 10^−2^ (Supplemental Table 1). Antibody responses to HCoV-NL63, HCoV-OC43 and MERS-CoV proteins were also among the thirty most discriminatory antigens for differentiating COVID-19 patients from control donors due to high reactivity with COVID-19 positive sera, while also demonstrating a considerable reactivity with negative. Nearly all the same epitopes and regions of reactivity found for IgG were recapitulated by IgA reactivity as well, when reactivity to the overlapping 100 aa, 50 aa and 30 aa protein fragments was analyzed (Fig. 3). This includes the epitopes mapped in the SARS-CoV-2 N, S1, S2 and M proteins (Fig. 2).

### Correlation of SARS-CoV-2 and endemic human coronavirus responses

By comparing the correlation between antibody responses to the S2 and N proteins of SARS-CoV-2 with responses to the S2 and N proteins of endemic human coronaviruses, in both COVID-19 positive and negative sera, we can estimate to what extent antibody responses to SARS-CoV-2 are the result of *de novo* immune responses or of boosting pre-existing immunity. There were significantly stronger correlations between SARS-CoV-2 S2 protein IgG and HCoV-OC43 S2 proteins in the positive group (Pearson’s correlation coefficient, *ρ*=0.6) than the negative group (*ρ*=0.24; Fig. 5A, top left). In the negative group, SARS-CoV-2 N protein IgG had no correlation with HCoV-OC43 N protein (*ρ*=0.02) or HCoV-NL63 N protein (*ρ*=0.09), whereas the correlations in the positive group were higher (HCoV-OC43 and HCoV-NL63 had *ρ*=0.44 with SARS-CoV-2 N protein). HCoV-OC43 and HCoV-NL63 N protein reactivity exhibited strong correlations in both positive and negative groups (*ρ*=0.54 and *ρ*=0.62, respectively). However, S2 protein reactivity correlations between these endemic human coronaviruses were lower in the negative group than the positive group (*ρ*=0.29 and *ρ*=0.49, respectively). Further inspection of the correlation scatterplot matrix (Supplemental Figure 3) showed a clear outlier in the CDC positive group for SARS-CoV-2 N protein and that correlations may differ if the CDC and Mayo cohorts were considered separately. Differential IgG reactivity between the COVID-19 positive and negative groups was also observed with the S2 and N proteins of SARS-CoV-2, HCoV-OC43 and HCoV-NL63. Positive COVID-19 patient sera had significantly higher IgG levels to S2 and N than the negative healthy donor sera for all three coronaviruses (Fig. 5B), with the exception of HCoV-OC43 N protein; this protein also showed higher IgA reactivity in the negatives (Supplemental Figure 4).

**Figure 5.**
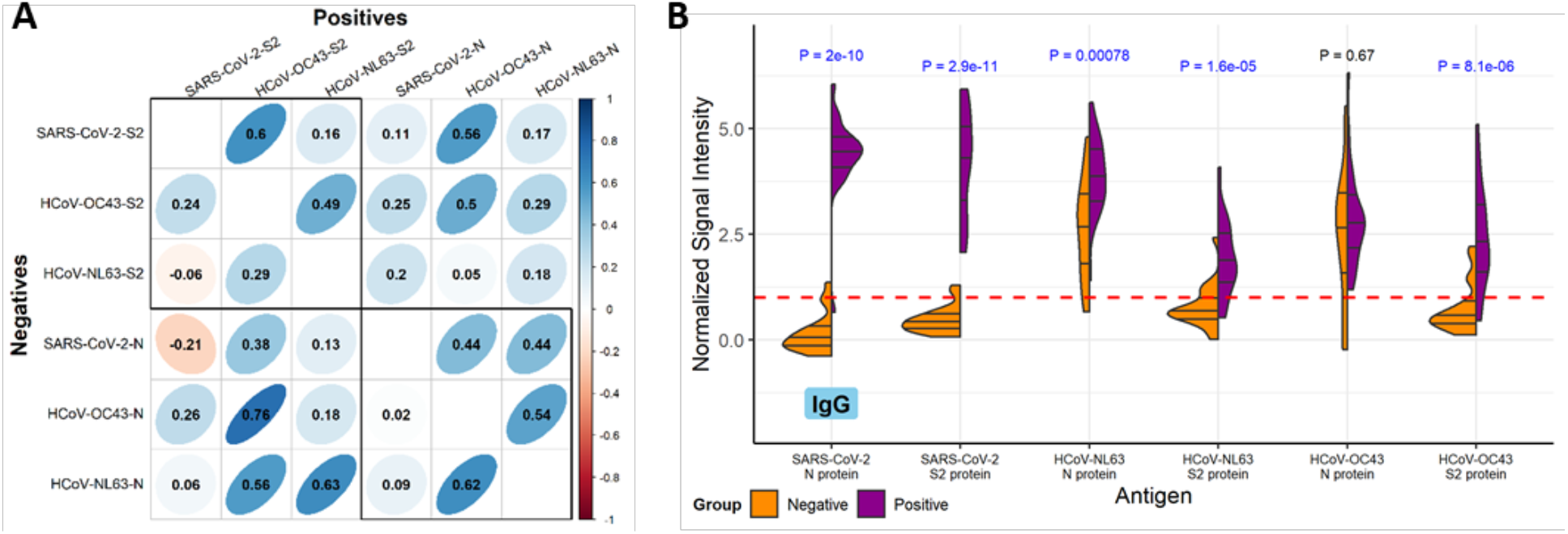
Correlation and concordance between IgG responses to SARS-CoV-2 and endemic human coronavirus N and S2 proteins. **(A)** The correlogram shows the Pearson’s correlation coefficient (*ρ*) between IgG normalized signal intensity to SARS-CoV-2, HCoV-OC43 and HCoV-NL63 N and S2 full-length proteins produced *in vitro*. The lower half of the diagonal shows correlation between reactivity of sera in the negative group, and the upper half of the diagonal shows the positive group serum correlations. The color scale indicates positive correlation in darker shades of blue and negative correlation in darker shades of red, and *ρ* is overlaid on each comparison. Additionally, the narrowness and slope of the ellipses represent increasing positive or negative correlation. Boxes are drawn around the intra-S2 and intra-N protein comparison. **(B)** The split violin plot shows the normalized IgG signal intensity distribution for each N and S2 protein produced *in vitro*. Within each half-violin are three lines representing the interquartile range and the median. Above each split violin is the Wilcoxon rank sum *p* value, colored blue for significant *p* values below 0.05. The red dashed line represents the 1.0 seropositivity cutoff.

### Correlation of multi-coronavirus protein microarray responses with ELISA and virus neutralization assays

S protein-based ELISA results from the CDC cohort, taken on all COVID-19 or healthy donor samples, were compared with IgG reactivity in the protein microarray data by Pearson’s correlation coefficient for the highly reactive IVTT S2 protein (*ρ*=0.85), IVTT N protein (*ρ*=0.9), purified recombinant full-length S protein (*ρ*=0.88) and the purified recombinant RBD (*ρ*=0.85), shown in Fig. 6A-D. The data clustered separately for negative responders and positive responders for all proteins. Virus neutralization titers were only available for the CDC COVID-19 patients and one healthy donor sample that tested near the 1.0 cutoff for ELISA reactivity (n=11). In all cases, neutralization activity was low, with positive neutralization titers at dilution factors of 20 or 40. Despite the low values and few samples, a trend was observed using linear regression for IVTT S2 (β=6.5, *p*=0.076), IVTT N (β=6, *p*=0.036) and stabilized purified S (β=6.3, *p*=0.077) (Fig. 6E-H). There was no association, however, of neutralization with responses to IgG reactivity with purified RBD (β=2.8, *p*=0.27). The linear regression models were specified with values of 0 for titers <20. However, since the true titer is between 0 and 20, neutralization was also modeled as an ordinal variable using ordinal logistic regression. Similar results were obtained for IVTT S2 and stabilized purified S, whereas association with IVTT N protein was no longer significant. The complete correlation results for all proteins are shown in Supplemental Table 1.

**Figure 6.**
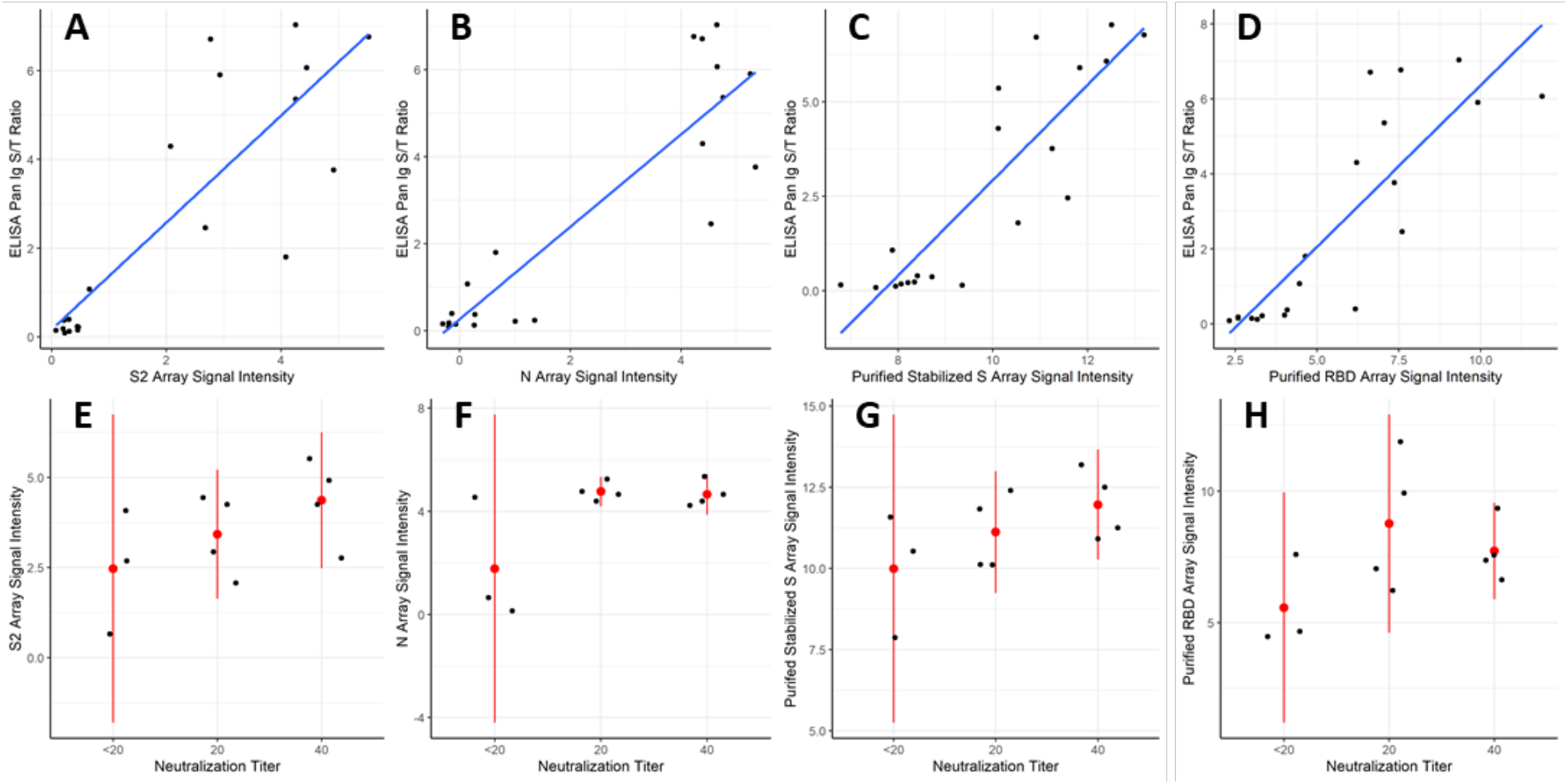
Correlation of IVTT and purified protein microarray results with ELISA and virus neutralization assays. (A-D) The scatter plots show the SARS-CoV-2 S protein-based ELISA Pan Ig signal/threshold ratio (y-axis) against the protein microarray normalized IgG signal intensity for S2 and N proteins produced in vitro, as well as for the stabilized, purified full-length S protein and the purified RBD fragment of S1 protein (x-axis), respectively. The blue lines were fit to the data using linear regression. (E-H) The dot plots show individual values of each patient for the protein microarray normalized IgG binding intensity (y-axis) of the four proteins shown in (A-D) at each neutralization titer (x-axis). Red dots are plotted at the means of each stratum, and the red lines represent the 95% confidence intervals.

## Discussion

In this study of twenty COVID-19 patients, the strongest antibody responses to the SARS-CoV-2 proteins used on this array, for all antibody isotypes, were directed to the N and S2 proteins as has been previously seen in other studies (5, 8, 14, 15). We also detected antibody responses to S1, M and accessory proteins 3a and 7a. Moreover, we were able to localize regions of each of these SARS-CoV-2 proteins to which antibodies bound, by antibody reactivity with overlapping protein fragments of three different lengths: 100, 50 and 30 aa. Our results were internally consistent in that reactive proteins had more reactive fragments than non-reactive proteins and 100 aa reactive fragments contained reactive 50 aa fragments and sometimes they also contained reactive 30 aa fragments. We found little reactivity of COVID-19 patient sera with 13-20 aa peptides from SARS-CoV S, M, E or N, HCoV-OC43 S or HCoV-NL63 S with the exception of one S2 peptide from HCoV-OC43.

Many previous publications have predicted B cell epitopes in SARS-CoV-2 proteins using a variety of immunoinformatic approaches (16-19). Crooke *et al*. predicted twenty-six potential linear B cell epitopes in the S protein, fourteen potential epitopes in the N protein, and three potential epitopes in the M protein. We noted antibody reactivity with regions containing some, but not all of these predicted epitopes. In particular, of the top six predicted B cell epitopes in the S protein we found significantly stronger reactivity with COVID-19 patient sera compared to healthy donor sera for regions containing three epitopes: DIADTT, residues 568-573 near the carboxy terminus of S1, PPIKD, residues 792-796 near the amino terminus of S2 and VYDPLQPELDSF, residues 1137-1148 near the carboxy terminus of S2. The other three top predicted B cell epitopes of the S protein, residues 405-428, 440-450 and 496-507, were not in highly reactive regions of the S protein in our experiments, perhaps due to the overall low reactivity of the S1 protein except for its carboxy terminal region or a need for native structure not found in protein fragments produced *in vitro*. Similarly, we found COVID-19 specific reactivity for regions including nine of the fourteen B cell epitopes in the N protein and one of three B cell epitopes in the M protein predicted by Crooke *et al*.

A few other groups have used protein or peptide arrays to map antibody reactivity to SARS-CoV-2 protein (15, 20-23). Two studies included full-length purified structural proteins from SARS-CoV-2, other human coronaviruses and diverse human retroviruses (15, 20). Their results are consistent with ours, but do not include accessory proteins or the ability to map reactive regions in each protein. Several groups used peptides to map epitopes in the SARS-CoV-2 S protein (21-23); Li *et al*. found four epitopes defined by 12 aa peptides, three of which are in regions of antibody reactivity that we found. Poh *et al*. found two epitopes defined by 18 aa peptides. Both are in regions of antibody reactivity that we described here. Finally, Zhang *et al*. used 15 aa peptides overlapping by 5 aa covering the whole SARS-CoV-2 proteome, plus full-length N and E as well as five truncated forms of S to map IgM and IgG responses of acute COVID-19 patients (median 4 days post onset of symptoms). They found a more robust IgM responses, though their specimens were collected earlier during infection. Zhang *et al*. identified five peptides as the most specific for COVID-19 patient IgG binding compared to controls: two in the S protein, two in N and one in ORF-1ab. Both S protein peptides are in regions where we found IgG reactivity; one is in the N-terminal region of reactivity we found in S2 and the other in the central reactivity region of S2. The N peptides of this group were not in a reactive region in our work and we did not assay antibody reactivity of the ORF-1ab polyproteins.

Recently two groups published epitope maps of SARS-CoV-2 using phage display (33, 34). One group analyzed 56 aa and 20 aa fragments of the SARS-CoV-2 proteome, while the other group analyzed 38 aa fragments of the proteome. Both studies also included other human coronaviruses and used COVID-19 patient sera and control sera to identify specifically reactive epitopes in the SARS-CoV-2 proteome. Their data are largely in agreement with data presented here. Both studies found the greatest reactivity of COVID-19 patient sera in the S2 and N proteins. Moreover, the epitopes they mapped overlapped with the ones we found here by different methods.

On the population level it is clear that groups of SARS-CoV-2 infected subjects have higher antibody levels to the whole N and S2 proteins, but it is also clear that even in the small sample sets evaluated here, some SARS-CoV-2 naïve individuals have substantial pre-existing antibody to some epitopes of these two proteins. Since the pre-existing antibody levels are likely to vary according to many different factors (e.g. geography, age, time of year and associated higher frequency of recent of exposure to other coronaviruses) it would be beneficial to have detailed knowledge of specific epitopes of the key immunogenic proteins.

The multi-coronavirus protein array is a tool that can help us improve our understanding of the immune response to SARS-CoV-2 and other coronaviruses. With these first two sets of convalescent sera provided by the Mayo Clinic and the CDC, we have shown that SARS-CoV-2 naïve subjects have clearly measurable cross-reactive antibody to the whole N and S2 proteins and that this reactivity is limited to specific epitopes. Importantly, there are epitopes that are more specific to SARS-CoV-2, that might serve as useful biomarkers of infection. Conversely, we have shown that infection with SARS-CoV-2 elicits or boosts the level of antibodies that bind to the N and S2 proteins of other coronaviruses including SARS-CoV, MERS-CoV, HCoV-NL63 and HCoV-OC43.

Limitations of our study are the small sample size and the inclusion of only convalescent samples. Despite these limitations, we were able to identify clear differences in the antibody response from COVID-19 patients and healthy, non-exposed controls. The ideal dataset to further investigate associations between preexisting antibody to specific epitopes and protection from severe disease would be longitudinal, with at least a pre-exposure sample, an acute sample, and a convalescent sample from each subject. Inclusion of samples from COVID-19 patients with a range of clinical symptoms will also provide an important comparison. In upcoming projects, we are seeking to analyze these types of samples paired with detailed clinical data on disease outcomes ranging from asymptomatic to fatal to further improve our understanding of the complex role antibodies play in SARS-CoV-2 infection. It may also prove interesting to test convalescent plasma samples, especially given the variable results on efficacy that have been reported in the literature (27-30). An assay providing more granular detail on the humoral response in these samples, such as the protein microarray described here, may provide valuable insights into what is happening during these convalescent plasma trials.

## Methods

### Protein microarray analysis of serum samples

The first generation multi-coronavirus protein microarray, produced by Antigen Discovery, Inc. (ADI, Irvine, CA, USA), included 935 full-length coronavirus proteins, overlapping 100, 50 and 30 aa protein fragments and overlapping 13-20 aa peptides from SARS-CoV-2 (WA-1), SARS-CoV, MERS-CoV, HCoV-NL63 and HCoV-OC43. Purified proteins and peptides were obtained from BEI Resources. All these coronavirus proteins were produced in *Escherichia coli* except the SARS-CoV-2 and SARS-CoV S proteins, which were made in Sf9 insect cells and the SARS-CoV-2 RBD, made in HEK-293 cells. Other proteins and protein fragments were expressed using an *E. coli in vitro* transcription and translation (IVTT) system (Rapid Translation System, Biotechrabbit, Berlin, Germany) and printed onto nitrocellulose-coated glass AVID slides (Grace Bio-Labs, Inc., Bend, OR, USA) using an Omni Grid Accent robotic microarray printer (Digilabs, Inc., Marlborough, MA, USA). Microarrays were probed with sera and antibody binding detected by incubation with fluorochrome-conjugated goat anti-human IgG or IgA or IgM (Jackson ImmunoResearch, West Grove, PA, USA or Bethyl Laboratories, Inc., Montgomery, TX, USA). Slides were scanned on a GenePix 4300A High-Resolution Microarray Scanner (Molecular Devices, Sunnyvale, CA, USA), and raw spot and local background fluorescence intensities, spot annotations and sample phenotypes were imported and merged in R (R Core Team, 2017), in which all subsequent procedures were performed. Foreground spot intensities were adjusted by subtraction of local background, and negative values were converted to one. All foreground values were transformed using the base two logarithm. The dataset was normalized to remove systematic effects by subtracting the median signal intensity of the IVTT controls for each sample. With the normalized data, a value of 0.0 means that the intensity is no different than the background, and a value of 1.0 indicates a doubling with respect to background. For full-length purified recombinant proteins and peptide libraries, the raw signal intensity data was transformed using the base two logarithm for analysis.

### Control Sera and COVID-19 Patient samples

COVID-19 positive and pre-COVID-19 negative control sera provided by the CDC were acquired from commercial laboratories or through partnership with Emory University. Samples were provided with only clinical and demographic information retained. The majority of samples (7/10) were from patients that were not hospitalized with blood collected between 26 and 60 days post symptom onset. Negative control sera were collected pre-COVID-19, in the fall of 2019. This activity was reviewed by CDC and was conducted consistent with applicable federal law and CDC policy (45 C.F.R. part 46, 21 C.F.R. part 56). The COVID-19 positive samples provided by Mayo Clinic were de-identified residual sera from clinical testing with only age and sex information available. The COVID-19 negative samples were collected pre-COVID-19 pandemic, between 2005-2012. These samples were from participants in prior Mayo Clinic vaccine studies who had provided informed consent for future use of their biospecimens. The original blood collection was collected through Mayo Clinic IRB-approved protocols. Samples were tested for SARS-CoV-2 specific antibodies and the presence of neutralizing antibodies as described below.

### Enzyme linked immunosorbent assay (ELISA)

CDC provided samples were tested using an enzyme-linked immunosorbent assay (ELISA) against the pre-fusion stabilized ectodomain of SARS-CoV-2 spike protein (31). This validated assay has been shown to have sensitivity and specificity of 96% and 99%, respectively (32). Briefly, plates were coated with purified spike protein and incubated overnight at 4°C followed by 37°C incubation steps and subsequent phosphate buffered saline + 0.05% Tween 20 (PBST) washings with: 2.5 X Stabilcoat blocker (Surmodics), 1:25 to 1:1600 diluted serum in 1 X PBST + 5% skim milk for 1 h, 1:2000 goat anti-human Ab conjugated to horseradish peroxidase (KPL) for 1h, ABTS peroxidase substrate for 30 min. Reactions were then quenched with stop solution. Plates were read at 405 nm and 490 nm, with resulting ODs calculated as 490 nm - 405 nm absorbance for each sample *and* minus PBS-only coated wells. Results are reported as a ratio of the calculated sample OD/cut-off threshold OD (signal/threshold, or S/T); values >1.0 are defined as positive. The Mayo Clinic COVID-19 positive samples were tested using an IgG SARS-CoV-2 Spike protein-specific ELISA assay (EuroImmune Inc.) performed according to manufacturer’s recommendations. This validated assay has been shown to have sensitivity and specificity of 90% and 100%, respectively (39). Results are reported as a ratio of the sample OD/calibrator OD (signal/calibrator, or S/C); values >1.1 are defined as positive.

### Neutralization assay

All SARS-CoV-2 microneutralization assays (MNT) were performed following biosafety level-3 precautions, using a SARS-CoV-2 clinical isolate. The WA1 strain of SARS-CoV-2 was employed using a modified version of a previously established protocol.27 Vero cell suspensions (ATCC CCL-81) were prepared at 2.2 – 2.5 × 105 cells/mL in DMEM (Thermo Fisher, catalog 11965118) + 10% fetal bovine serum (FBS, defined, Hyclone catalog SH30070.03, heat-inactivated 56°C for 30 min) + 2X antibiotic-antimycotic (Thermo Fisher catalog 15240062) + 2X penicillin-streptomycin (Thermo Fisher catalog 15140122) immediately before use. Sera were 2-fold serial diluted in serum-free DMEM in a 96-well flat bottom plate, from 1:10 – 1:320, in triplicate, to a final volume of 50 µL/well. Fifty µL SARS-CoV-2 was to each well, such that final serum dilution titers ranged from 1:20 – 1:640. After 30 min incubation at 37°C and 5% CO2, 100 µL of Vero cells in suspension were added to each well, for a final concentration of 2.2 – 2.5 × 104 cells/well. After 5 days cells were stained and fixed with crystal violet fixative (0.15% crystal violet, 2.5% ethanol, 11% formaldehyde, 50% PBS, 0.01M pH 7.4). The endpoint concentration at which antibodies were determined to be neutralizing for SARS-CoV-2 infection was the lowest concentration of antibody at which 3 replicate wells were protected against virus infection.

### Statistical Analysis

Student’s t-tests were used for comparison of the individual antibody response means between the positive and negative groups. Comparison of the medians was done using Wilcoxon’s rank sum test. The area under the receiver operating characteristics curve (AUC) was calculated to estimate delineation of groups for each antigen. The t-SNE analyses were calculated after 25,000 iterations with a perplexity parameter of 30 using the R package Rtsne (35). Comparisons of the proportions of responders to each protein between groups was done using two-proportions z-tests implemented by the ‘prop.test’ function in R. Correlation between antibody features and between protein microarray and ELISA measurements used Pearson’s correlation coefficient (*ρ*), and association between antibody measurements and sample information such as sex, age and cohort were modeled using linear regression. The association of specific antibody responses with virus neutralization titers was estimated using linear regression with the values below detection levels (<20) coded as zero, or by converting neutralization titers to ordinal values and estimating the proportional odds ratio by ordinal logistic regession, whereby *p* values were estimated by comparing the t-value against the standard normal distribution. Adjustment for the false discovery rate was performed using the “p.adjust” function in R (36). Data visualization was performed using the circlize (37), ComplexHeatmap (38), ggplot2, heatmap2 and corrplot (39) packages in R. Unadjusted *p* values were shown in graphics.

## Data Availability

All data are freely available.

## Acknowledgements

We thank the Laboratory Task Force of the CDC COVID_19 response for their project review and resource support. This research was made possible using samples obtained from the CDC Biorepository.

The findings and conclusions in this report are those of the author(s) and do not necessarily represent the official position of the Centers for Disease Control and Prevention. Names of specific vendors, manufacturers, or products are included for public health and informational purposes; inclusion does not imply endorsement of the vendors, manufacturers, or products by the Centers for Disease Control and Prevention or the US Department of Health and Human Services.

## Author Contributions

DC, AY, XL, JJC, AZR, AO and AAT designed the Multi-coronavirus microarray; AY, XL, JJC, MT, SP, RK designed the study, arranged sample selection and testing; AAT, CH, JVP, JE, VH and AS fabricated the array and performed experiments; MS, SNL,JH, AT, MR, and NJT designed and performed ELISA and neutralization assay experiments. AZR, JJC and AO performed statistical analysis and data visualization; and DC, AZR, JJC, RK, SP, MT and AY wrote and reviewed the manuscript.

## References

1. https://www.who.int/news-room/detail/08-04-2020-who-timeline---covid-19

2. https://coronavirus.jhu.edu/map.html

3. Kissler S, Tedijanto C, Goldstein E, Grad Y and Lipsitch M (2020) Projecting transmission dynamics of SARS-CoV-2 through the postpandemic period. Science 10.1126/science.abb5793

4. Verity R et al. (2020) Estimates of the severity of coronavirus disease 2019: a model-based analysis. Lancet Infect Dis https://doi.org/10.1016/S1473-3099(20)30243-7

5. Long, Antibody responses to SARS-CoV-2 in patients with COVID-19, 2020

6. Zhao, Antibody responses to SARS-CoV-2 in patients of novel coronavirus disease 2019, 2020

7. Yu, Distinct features of SARS-CoV-2-specific IgA response in COVID-19 patients, 2020

8. Guo, Profiling Early Humoral Response to Diagnose Novel Coronavirus Disease (COVID-19), 2020

9. Atyeo, Distinct Early Serological Signatures Track with SARS-CoV-2 Survival, 2020

10. McAndrews, Heterogeneous antibodies against SARS-CoV-2 spike receptor binding domain and nucleocapsid with implications on COVID-19 immunity, 2020

11. Yang, Evasion of antibody neutralization in emerging severe acute respiratory syndrome coronaviruses, 2005

12. Jaume-Anti-Severe Acute Respiratory Syndrome Coronavirus Spike Antibodies Trigger Infection of Human Immune Cells via a pH- and Cysteine Protease-Independent FcgammaR Pathway, 2011

13. Yip, Antibody-dependent infection of human macrophages by severe acute respiratory syndrome coronavirus 2014

14. Brouwer et al., Science 369, 643–650 (2020)

15. de Assis R et al., Analysis of SARS-CoV-2 Antibodies in COVID-19 Convalescent Plasma using a Coronavirus Antigen Microarray, https://doi.org/10.1101/2020.04.15.043364 (2020)

16. Crooke et al., https://doi.org/10.1038/s41598-020-70864-8 (2020)

17. Grifoni et al., Cell Host & Microbe 27, 671–680, https://doi.org/10.1016/j.chom.2020.03.002 (2020)

18. Feng, Y.-E. et al.. Multi-epitope vaccine design using an immunoinformatics approach for 2019 novel coronavirus in China, https://doi.org/10.1101/2020.03.03.962332 (2020)

19. Fast E and Chen B, Potential T-cell and B-cell Epitopes of 2019-nCoV, https://doi.org/10.1101/2020.02.19.955484 (2020)

20. Khan S et al., Analysis of Serologic Cross-Reactivity Between Common Human Coronaviruses and SARS-CoV-2 Using Coronavirus Antigen Microarray, https://doi.org/10.1101/2020.03.24.006544 (2020)

21. Li, Linear epitopes of SARS-CoV-2 spike protein elicit neutralizing antibodies in COVID-19 patients, https://doi.org/10.1038/s41423-020-00523-5 (2020)

22. Poh C, Potent neutralizing antibodies in the sera of 1 convalescent COVID-19 patients 2 are directed against conserved linear epitopes on the SARS-CoV-2 spike 3 protein, https://doi.org/10.1101/2020.03.30.015461 (2020)

23. Zhang X, Proteome-wide analysis of differentially-expressed SARS-CoV-2 antibodies in early COVID-19 infection, https://doi.org/10.1101/2020.04.14.20064535 (2020)

24. Wang Q et al., Immunodominant SARS Coronavirus Epitopes in Humans Elicited both Enhancing and Neutralizing Effects on Infection in Non-human Primates, DOI: 10.1021/acsinfecdis.6b00006 (2016)

25. Chandrashekar et al., SARS-CoV-2 infection protects against rechallenge in rhesus macaques. Science 369 (6505), 812–817. DOI: 10.1126/science.abc4776 (2020)

26. Deng et al., Primary exposure to SARS-CoV-2 protects against reinfection in rhesus macaques. Science 369 (6505), 818–823. DOI: 10.1126/science.abc5343 (2020)

27. Liu et al., Convalescent plasma treatment of severe COVID-19: a propensity score–matched control study, Nat Med. doi: 10.1038/s41591-020-1088-9. (2020)

28. Wang et al., Evaluation of current medical approaches for COVID-19: a systematic review and meta-analysis. doi:10.1136/bmjspcare-2020-002554 (2020)

29. Bakhatawar et al., Convalescent Plasma Therapy and Its Effects On COVID-19 Patient Outcomes: A Systematic Review of Current Literature. Cureus 12(8): e9535. DOI 10.7759/cureus.9535 (2020)

30. Wooding and Bach, Treatment of COVID-19 with convalescent plasma: lessons from pastcoronavirus outbreaks. Clin Microbiol Infect. 2020 26:1436–1446. https://doi.org/10.1016/j.cmi.2020.08.005. (2020)

31. Wrapp D, Wang N, Corbett KS, Goldsmith JA, Hsieh CL, Abiona O, et al. Cryo-EM structure of the 2019-nCoV spike in the prefusion conformation. Science. 2020;367:1260–3.

32. Freeman B, Lester S, Mills L, et al. Validation of a SARS-CoV-2 spike protein ELISA for use in contact investigations and serosurveillance. bioRxiv. 2020 Apr 25; 2020.04.24.057323. doi: 10.1101/2020.04.24.057323. Preprint. https://www.biorxiv.org/content/10.1101/2020.04.24.057323v2.full

33. Shrock E et al. Viral epitope profiling of COVID-19 patients reveals cross-reactivity and correlates of severity. Science 10.1126/science.abd4250 (2020).

34. Zamecnik, C et al.. ReScan, a Multiplex Diagnostic Pipeline, Pans Human Sera for SARS-CoV-2 Antigens. https://doi.org/10.1016/j.xcrm.2020.100123 (2020)

35. Benjamini, Y., and Hochberg, Y. (1995). Controlling the false discovery rate: a practical and powerful approach to multiple testing. Journal of the Royal Statistical Society Series B, 57, 289–300. http://www.jstor.org/stable/2346101.

36. Gu, Z. (2014) circlize implements and enhances circular visualization in R. Bioinformatics. DOI: 10.1093/bioinformatics/btu393

37. Gu, Z. (2016) Complex heatmaps reveal patterns and correlations in multidimensional genomic data. DOI: 10.1093/bioinformatics/btw313

38. Michael Friendly (2002). Corrgrams: Exploratory displays for correlation matrices. The American Statistician, 56, 316–324.

39. https://www.fda.gov/medical-devices/coronavirus-disease-2019-covid-19-emergency-use-authorizations-medical-devices/eua-authorized-serology-test-performance

