## Supplement for "Mapping SARS-CoV-2 Antibody Epitopes in COVID-19 Patients with a Multi-Coronavirus Protein Microarray"

### Supplemental Figures

David Camerini[1,2], Arlo Z. Randall[1], Krista Trappl-Kimmons[1], Amit Oberai[1], Christopher Hung[1], Joshua Edgar[1], Adam Shandling[1], Vu Huynh[1], Andy A. Teng[1], Gary Hermanson[1], Jozelyn V. Pablo[1], Megan M. Stumpf[3], Sandra N. Lester[3], Jennifer Harcourt[3], Azaibi Tamin[3], Mohammed Rasheed[3], Natalie J. Thornburg[3], Panayampalli S. Satheshkumar[3], Xiaowu Liang[1], Richard B. Kennedy[4], Angela Yee[1] Michael Townsend[3*], and Joseph J. Campo[1*]

[1]Antigen Discovery Incorporated, Irvine CA; [2]University of California, Irvine, CA; [3]Centers for Disease Control and Prevention, Atlanta, GA; [4]Mayo Clinic, Rochester, MN; *co-senior authors.

**Figure S1**

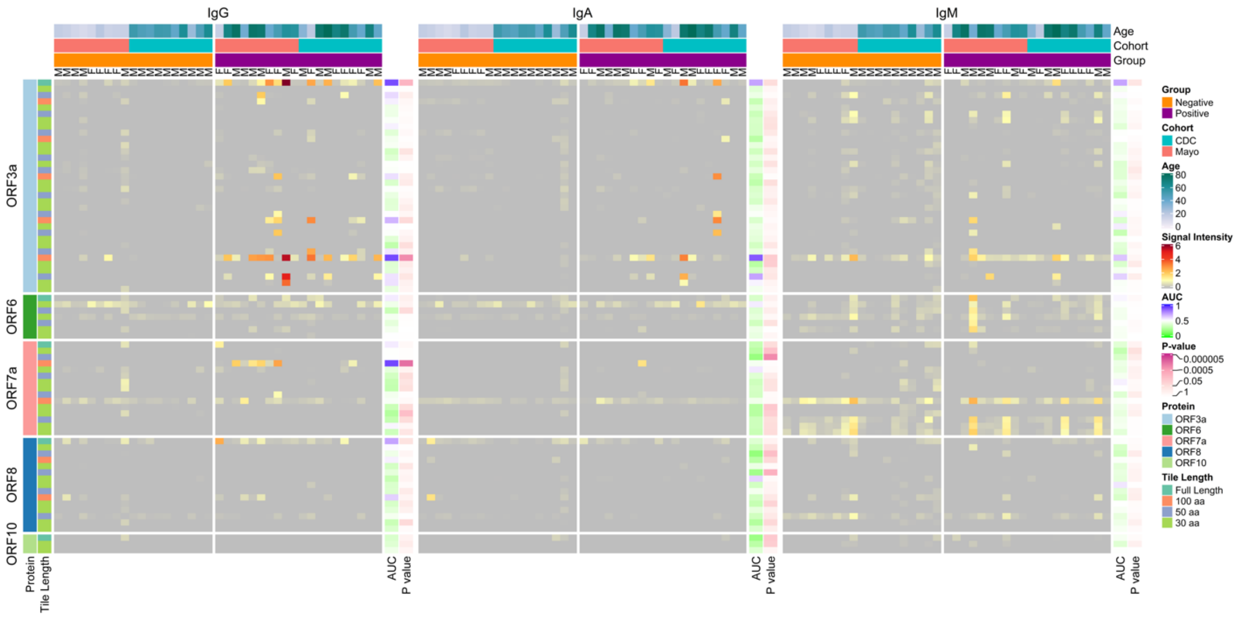


**Figure S1. Reactivity in COVID-19 patient and healthy donor IgG (left heatmap), IgA (middle heatmap) and IgM (right heatmap) to SARS-CoV-2 nonstructural proteins and protein fragments produced *in vitro*.** The heatmaps present the signals of individual array spots for the nonstructural SARS-CoV-2 proteins produced by IVTT. Columns represent serum samples, and rows represent IVTT proteins and protein fragments (n=34 ORF3a, n=7 ORF6, n=15 ORF7a, n=15 ORF8, n=3 ORF10). Antibody signal intensity is shown on a color scale from grey to red. Sample information is overlaid above the heatmaps and includes sex (M/F), group (Negative or Positive), cohort (CDC or Mayo) and age (years). Protein/fragment information is annotated to the left of the heatmaps and includes the full-length protein name and the amino acid length of the protein fragments (“Tile Length”, as full length, 100, 50 or 30 aa). Proteins are ordered by the starting amino acid position for each fragment. For each isotype, the receiver operating characteristic area under the curve (AUC) and the unadjusted t-test *p* value between negatives and positives are shown to the right of each heatmap.

**Figure S2**

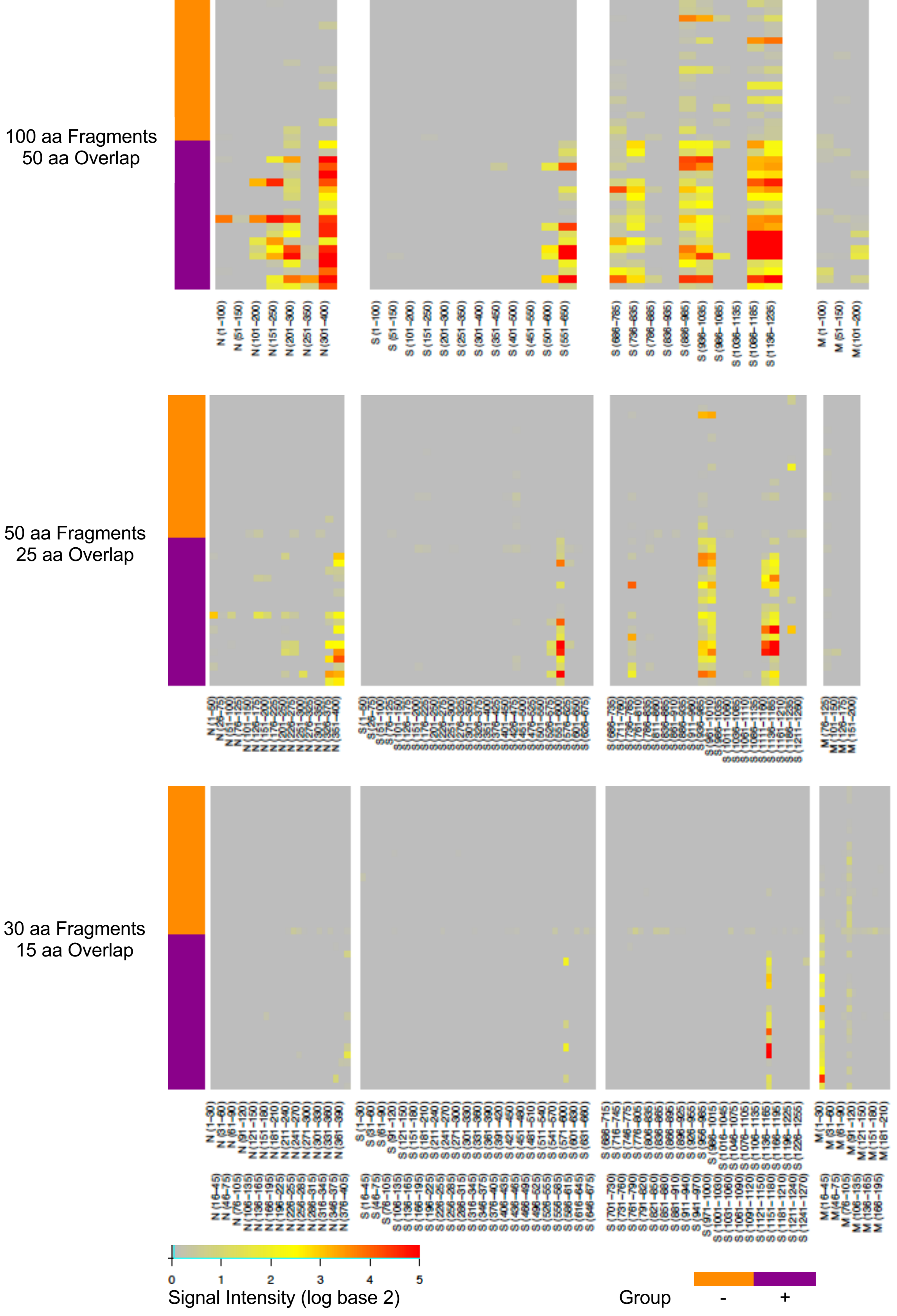
N
S1
S2
M
100 aa Fragments
50 aa Overlap
50 aa Fragments
25 aa Overlap
30 aa Fragments
15 aa Overlap
0 1 2 3 4 5
Signal Intensity (log base 2)
Group
-
+

**Figure S2. COVID-19 patient IgG reactivity with SARS-CoV-2 N, S and M protein fragments arranged in amino acid sequence order.** Heat maps show the IgG reactivity of each serum sample separately in each row. Columns denote each protein fragment as labeled; the 30 aa fragment labels are staggered to save space. A scale shows the meaning of the colors. Bars at left of each heat map identify the samples: orange indicates negative control samples and purple shows COVID-19 positive samples. Amino acid numbers indicate the positions of the fragments in each protein.

**Figure S3**

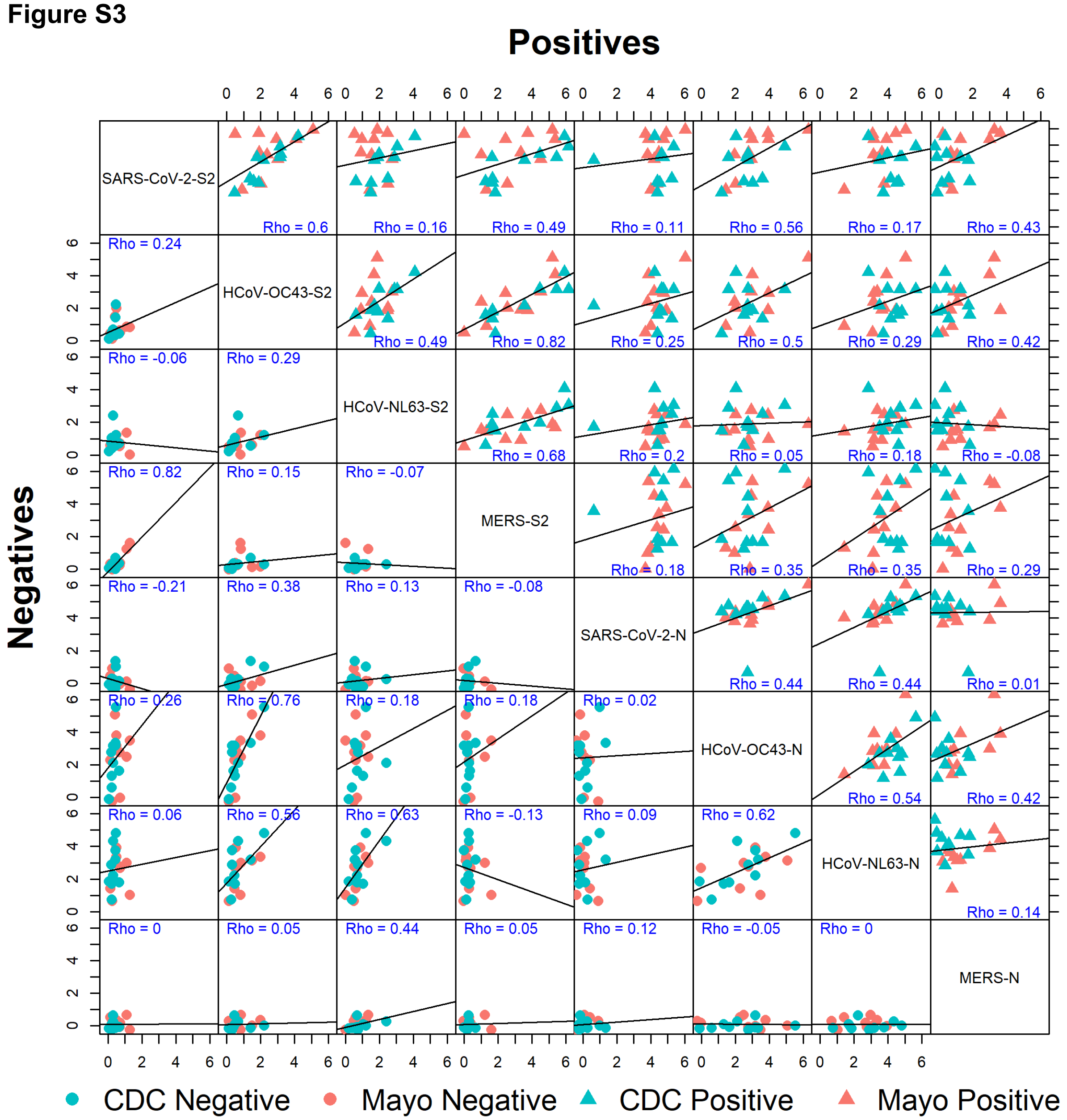


**Figure S3. Correlation between IgG responses to SARS-CoV-2 and other human coronavirus N and S2 proteins.** The correlation matrix shows the Pearson's correlation coefficient ("Rho", blue) between IgG normalized signal intensity to SARS-CoV-2, MERS-CoV, HCoV-OC43 and HCoV-NL63 N and S2 full-length proteins produced *in vitro*. The lower half of the diagonal shows correlation between proteins in the negative group (circles), and the upper half of the diagonal shows the positive group correlations (triangles). Rho values and a linear regression line are overlaid on each comparison.

**Figure S4**

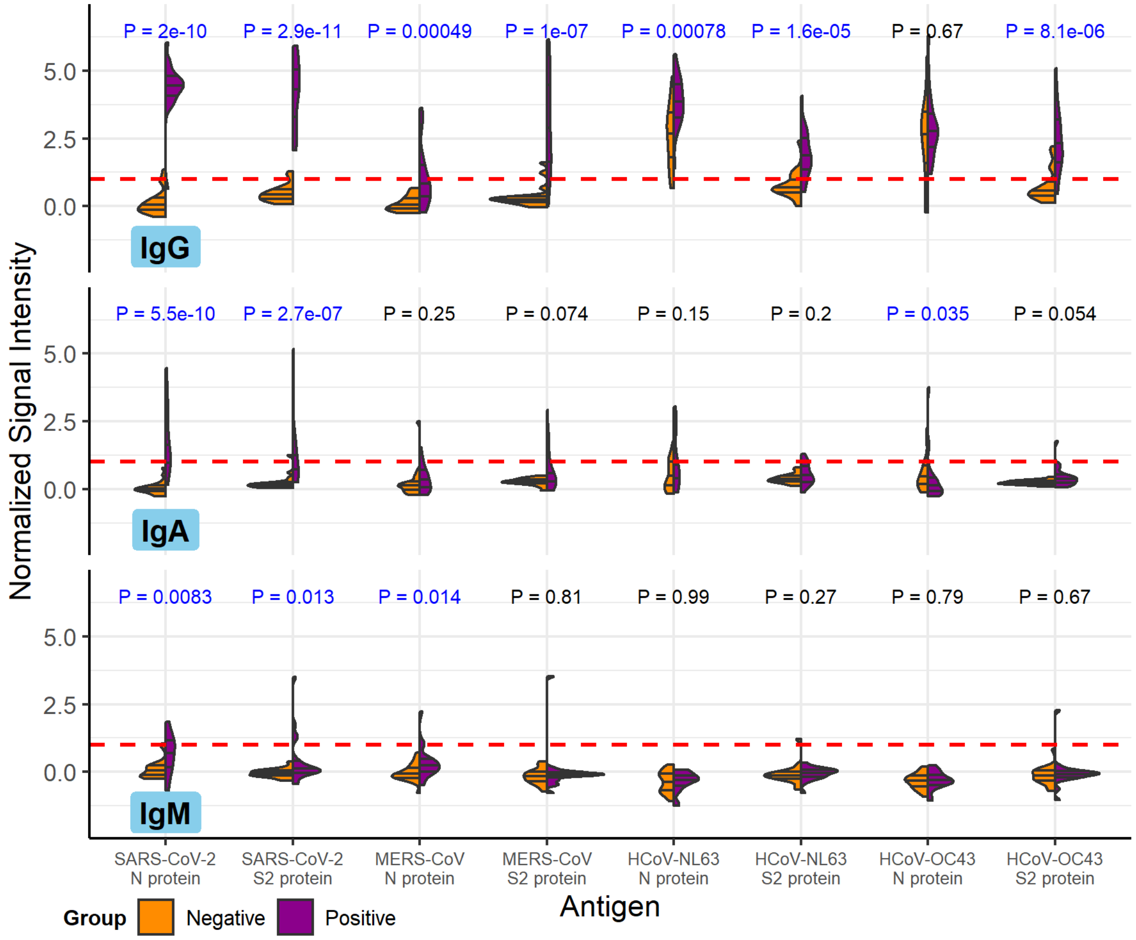


**Figure S4. Differential reactivity between IgG, IgA and IgM responses to SARS-CoV-2 and other human coronavirus N and S2 proteins.** The results show the differential reactivity between IgG, IgA and IgM normalized signal intensity to SARS-CoV-2, MERS-CoV, HCoV-OC43 and HCoV-NL63 N and S2 full-length proteins produced *in vitro* on a log scale, base 2. The split violin plot shows the normalized signal intensity distribution of each of the IVTT N and S2 proteins. Within each half-violin are three lines representing the interquartile range and the median. Above each split violin is the Wilcoxon rank sum *p* value, colored blue for significant *p* values below 0.05. The red dashed line represents the 1.0 seropositivity cutoff. The healthy control negative group is shown on the left violin halves in orange, and the COVID-19 patient positive group is shown in the right violin halves in purple.
